# Lopinavir/ritonavir for the treatment of COVID-19: A living systematic review protocol

**DOI:** 10.1101/2020.04.11.20062109

**Authors:** Francisca Verdugo-Paiva, Ariel Izcovich, Martín Ragusa, Gabriel Rada

**Affiliations:** Fundación Epistemonikos, Santiago, Chile; UC Evidence Center, Cochrane Chile Associated Center, Pontificia Universidad Católica de Chile, Santiago, Chile; Internal Medicine Service, German Hospital, Buenos Aires, Argentina; Internal Medicine Department, Faculty of Medicine, Pontificia Universidad Católica de Chile, Santiago, Chile

**Keywords:** COVID-19, severe acute respiratory syndrome coronavirus 2, Coronavirus Infections, Systematic review, lopinavir, lopinavir/ritonavir, antivirals

## Abstract

**Objective:** To assess the efficacy and safety of lopinavir/ritonavir for the treatment of patients with COVID-19.

**Design:** This is the protocol of a living systematic review.

**Data sources:** We will conduct searches in PubMed/Medline, Embase, Cochrane Central Register of Controlled Trials (CENTRAL), grey literature and in a centralised repository in L·OVE (Living OVerview of Evidence). L·OVE is a platform that maps PICO questions to evidence from Epistemonikos database. In response to the COVID-19 emergency, L·OVE was adapted to expand the range of evidence it covers and customised to group all COVID-19 evidence in one place. The search will cover the period until the day before submission to a journal.

**Eligibility criteria for selecting studies and methods:** We adapted an already published common protocol for multiple parallel systematic reviews to the specificities of this question.

We will include randomised trials evaluating the effect of lopinavir/ritonavir— as monotherapy or in combination with other drugs — versus placebo or no treatment in patients with COVID-19. Randomised trials evaluating lopinavir/ritonavir in infections caused by other coronaviruses, such as MERS-CoV and SARS-CoV, and non-randomised studies in COVID-19 will be searched in case no direct evidence from randomised trials is found, or if the direct evidence provides low- or very low-certainty for critical outcomes.

Two reviewers will independently screen each study for eligibility, extract data, and assess the risk of bias. We will perform random-effects meta-analyses and use GRADE to assess the certainty of the evidence for each outcome.

A living, web-based version of this review will be openly available during the COVID-19 pandemic. We will resubmit it if the conclusions change or there are substantial updates.

**Ethics and dissemination:** No ethics approval is considered necessary. The results of this review will be widely disseminated via peer-reviewed publications, social networks and traditional media.

**PROSPERO Registration:** Submitted to PROSPERO (awaiting ID allocation).

## INTRODUCTION

COVID-19 is an infection caused by the SARS-CoV-2 coronavirus [1], first identified in Wuhan, China, on December 31, 2019 [2]. Three months later, almost half a million cases of contagion had been identified across 197 countries [3], and on March 11 2020 the WHO characterised the COVID-19 outbreak as a pandemic [1].

While the majority of cases result in mild symptoms, some might progress to pneumonia, acute respiratory distress syndrome and death [4],[5],[6]. The case fatality rate reported across countries, settings and age groups is highly variable, but it would range from about 0.5% to 10% [7]. In hospitalised patients it has been reported to be higher than 10% in some centres [8]. Consequently, identifying treatment options is critical for the response to the COVID-19 outbreak.

Lopinavir/ritonavir is a fixed-dose combination antiviral widely used for HIV infection and it has been suggested as a possible treatment in the context of the COVID-19 pandemic in 2019/2020. The information about its antiviral properties against coronavirus comes from its use in the previous SARS-CoV and MERS-CoV pandemics. In vitro studies demonstrated an antiviral activity of lopinavir/ritonavir against SARS-CoV [9] and clinical studies reported a reduction in the intubation rate, steroid requirements and mortality with lopinavir/ritonavir in SARS patients [10].

Notwithstanding, a number of systematic reviews have evaluated the role of lopinavir/ritonavir concluding it was not possible to establish its efficacy, based on the results of non-randomised trials [11] [12] [13].

In the last few weeks, multiple reviews assessing the role of this treatment for COVID-19 have been published [14] [15] [16]. However, due to the high speed at which new primary studies are being published, they are all missing relevant evidence.

Using innovative and agile processes, taking advantage of technological tools, and resorting to the collective effort of several research groups, this living systematic review aims to provide a timely, rigorous and continuously updated summary of the evidence available on the in patients with COVID-19.

## METHODS

### Protocol and registration

This manuscript complies with the ‘Preferred Reporting Items for Systematic reviews and Meta-Analyses’ (PRISMA) guidelines for reporting systematic reviews and meta-analyses [17].

A protocol stating the shared objectives and methodology of multiple systematic reviews to be conducted in parallel for different questions relevant to COVID-19 was published elsewhere [18]. This protocol was adapted to the specificities of the question assessed in this review and submitted to PROSPERO (awaiting ID allocation)

### Search strategies

#### Electronic searches

Our literature search was devised by the team maintaining the L·OVE platform (https://app.iloveevidence.com), using the following approach:

1. Identification of terms relevant to the population and intervention components of the search strategy, using Word2vec technology [19] to the corpus of documents available in Epistemonikos Database.
2. Discussion of terms with content and methods experts to identify relevant, irrelevant and missing terms.
3. Creation of a sensitive boolean strategy encompassing all of the relevant terms
4. Iterative analysis of articles missed by the boolean strategy, and refinement of the strategy accordingly.

Our main search source was Epistemonikos database (https://www.epistemonikos.org), a comprehensive database of systematic reviews and other types of evidence [20]. We supplemented it with articles from multiple sources relevant to COVID-19 (without any study design, publication status or language restriction) [21].

In sum, Epistemonikos Database acts as a central repository. Only articles fulfilling Epistemonikos criteria are visible by users. The remaining articles are only accessible for members of COVID-19 L·OVE Working Group.

Additional searches will be conducted using highly sensitive searches in PubMed/MEDLINE, the Cochrane Central Register of Controlled Trials (CENTRAL), Embase and the WHO International Clinical Trials Registry Platform, without any language or publication status restriction. The searches will cover from the inception date of each database until the day before submission.

The following strategy will be used to search in Epistemonikos Database. We will adapt it to the syntax of other databases.

(“covid-19” OR covid19* OR “covid 19” OR “2019-nCoV” OR cv19* OR “cv-19” OR “cv 19” OR “n-cov” OR ncov* OR “sars-cov-2” OR “sars-cov2” OR “2019-ncov” OR “SARS-Coronavirus-2” OR “SARS-Coronavirus2” OR (wuhan* AND (virus OR viruses OR viral)) OR (covid* AND (virus OR viruses OR viral)) OR “covid-19-related” OR “SARS-CoV-2-related” OR “SARS-CoV2-related” OR “2019-nCoV-related” OR “cv-19-related” OR “n-cov-related”) AND (lopinavir* OR “ABT-378” OR “ABT 378” OR ABT378* OR Kaletra* OR ritonavir* OR Norvir)

#### Other sources

In order to identify articles that might have been missed in the electronic searches, we will do the following:

1. Screen the reference lists of other systematic reviews, and evaluate in full text all the articles they include.
2. Scan the reference lists of selected guidelines, narrative reviews and other documents.
3. Conduct cross-citation search in Google Scholar and Microsoft Academic, using each included study as the index reference.
4. Review websites from pharmaceutical companies producing drugs claimed as effective for COVID-19, websites or databases of major regulatory agencies, and other websites specialised in COVID-19.
5. Email the contact authors of all of the included studies to ask for additional publications or data on their studies, and for other studies in the topic.
6. Review the reference list of each included study.

### Eligibility criteria

#### Types of studies

We will preferentially include randomised trials. However, information from non-randomised studies will be used if there is no direct evidence from randomised trials or the certainty of evidence for the critical outcomes resulting from the randomised trials is graded as low- or very low, and the certainty provided by the non-randomised evidence grades higher than the one provided by the randomised evidence [22].

Studies evaluating the effects on animal models or in vitro conditions will be excluded.

#### Types of participants

We will include trials assessing participants with COVID-19, as defined by the authors of the trials.

If substantial clinical heterogeneity on how the condition was defined is found, we will explore it using a sensitivity analysis.

In case no direct evidence from randomised trials is found, or if the evidence from randomised trials provides low- or very low-certainty evidence for critical outcomes, we will include information from randomised trials evaluating lopinavir/ritonavir in other coronavirus infections (i.e. MERS-CoV or SARS-CoV) [22].

#### Type of interventions

The intervention of interest is the combination of lopinavir/ritonavir. We will not restrict our criteria to any dosage, duration, timing or route of administration.

The comparison of interest will be placebo (lopinavir/ritonavir plus optimal treatment versus placebo plus optimal treatment) or no treatment (lopinavir/ritonavir plus optimal treatment versus optimal treatment).

Trials assessing lopinavir/ritonavir plus other drugs will be eligible if the cointerventions are identical in both intervention and comparison groups.

Trials evaluating lopinavir/ritonavir in combination with other active drugs versus placebo or no treatment will be also included.

#### Type of outcomes

We will not use the outcomes as an inclusion criteria during the selection process. Any article meeting all the criteria except for the outcome criterion will be preliminarily included and evaluated in full text. We used the core outcome set COS-COVID [23], the existing guidelines and reviews and the judgement of the authors of this review as an input to select the primary and secondary outcomes, as well as to decide upon inclusion. The review team will revise this list of outcomes, in order to incorporate ongoing efforts to define Core Outcomes Sets (e.g. COVID-19 Core Outcomes [24].

##### Primary outcome

- All-cause mortality

##### Secondary outcomes

- Mechanical ventilation
- Extracorporeal membrane oxygenation
- Length of hospital stay
- Respiratory failure
- Serious adverse events
- Time to SARS-CoV-2 RT-PCR negativity

##### Other outcomes

- Acute respiratory distress syndrome
- Total adverse effects

Primary and secondary outcomes will be presented in the GRADE ‘Summary of Findings’ tables, and a table with all the outcomes will be presented as an appendix [25].

### Selection of studies

The results of the literature search in Epistemonikos database will be automatically incorporated into the L·OVE platform (automated retrieval), where they will be de-duplicated by an algorithm comparing unique identifiers (database ID, DOI, trial registry ID), and citation details (i.e. author names, journal, year of publication, volume, number, pages, article title and article abstract).

The additional searches will be uploaded to the screening software Collaboratron(tm) [26].

In both L·OVE platform and Collaboratron(tm), two researchers will independently screen the titles and abstracts yielded by the search against the inclusion criteria. We will obtain the full reports for all titles that appear to meet the inclusion criteria or require further analysis to decide on their inclusion.

We will record the reasons for excluding trials in any stage of the search and outline the study selection process in a PRISMA flow diagram adapted for the purpose of this project.

### Extraction and management of data

Using standardised forms, two reviewers will extract data independently from each included study. We will collect the following information: study design, setting, participant characteristics (including disease severity and age) and study eligibility criteria; details about the administered intervention and comparison, including dose and therapeutic scheme, duration, timing (i.e. time after diagnosis) and route of administration; the outcomes assessed and the time they were measured; the source of funding of the study and the conflicts of interest disclosed by the investigators; the risk of bias assessment for each individual study.

We will resolve disagreements by discussion, and one arbiter will adjudicate unresolved disagreements

### Risk of bias assessment

The risk of bias for each randomised trial will be assessed using the ‘risk of bias’ tool (RoB 2.0: a revised tool to assess risk of bias in randomised trials) [27]. We will consider the effect of assignment to the intervention for this review. Two reviewers will independently assess five domains of bias for each outcome result of all reported outcomes and time points. These five domains are: bias due to (1) the randomisation process, (2) deviations from intended interventions (effects of assignment to interventions at baseline), (3) missing outcome data, (4) measurement of the outcome, and (5) selection of reported results. Answers to signalling questions and supporting information collectively will lead to a domain-level judgement in the form of ‘Low risk of bias’, ‘Some concerns’, or ‘High risk of bias’. These domain-level judgements will inform an overall ‘risk of bias’ judgement for each result. Discrepancies between review authors will be resolved by discussion to reach consensus. If necessary, a third review author will be consulted to achieve a decision.

We will assess the risks of bias of other study designs with the ROBINS-I tool (ROBINS-I: Risk Of Bias In Non-randomised Studies of Interventions) [28]. We will assess the following domains: bias due to confounding, bias in selection of participants into the study, bias in classification of interventions, bias due to deviations from intended interventions (effect of assignment to intervention), bias due to missing data, bias in measurement of outcomes and bias in the selection of the reported result. We will judge each domain as low risk, moderate risk, serious risk, critical risk, or no information, and evaluate individual bias items as described in ROBINS-I guidance. We will not consider time-varying confounding, as these confounders are not relevant in this setting [28]. We will consider the following factors as baseline potential confounders:

- Age
- Comorbidity (e.g. cardiovascular disease, renal disease, liver disease)
- Co-interventions
- Severity, as defined by the authors (i.e respiratory failure vs respiratory distress syndrome vs ICU requirement).

### Measures of treatment effect

For dichotomous outcomes, we will express the estimate of treatment effect of an intervention as risk ratios (RR) or odds ratios (OR) along with 95% confidence intervals (CI).

For continuous outcomes, we will use mean difference and standard deviation (SD) to summarise the data using a 95% CI. Whenever continuous outcomes are measured using different scales, the treatment effect will be expressed as a standardised mean difference (SMD) with 95% CI. When possible, we will multiply the SMD by a standard deviation that is representative from the pooled studies, for example, the SD from a well-known scale used by several of the studies included in the analysis on which the result is based. In cases where the minimally important difference (MID) is known, we will also present continuous outcomes as MID units or inform the results as the difference in the proportion of patients achieving a minimal important effect between intervention and control [29].

Then, these results will be displayed on the ‘Summary of Findings Table’ as mean difference [29].

### Strategy for data synthesis

If we include more than one trial we will conduct meta-analysis for studies clinically homogeneous using RevMan 5 [30], using the inverse variance method with random effects model. For any outcomes where data was insufficient to calculate an effect estimate, a narrative synthesis will be presented.

### Subgroup and sensitivity analysis

We will perform subgroup analysis according to the definition of severe COVID-19 infection (i.e respiratory failure vs respiratory distress syndrome vs ICU requirement). In case we identify significant differences between subgroups (test for interaction <0.05) we will report the results of individual subgroups separately.

We will perform sensitivity analysis excluding high risk of bias studies, and if non-randomised studies are used, excluding studies that did not report adjusted estimates. In cases where the primary analysis effect estimates and the sensitivity analysis effect estimates significantly differ we will either present the low risk of bias — adjusted sensitivity analysis estimates — or present the primary analysis estimates but downgrading the certainty of the evidence because of risk of bias.

### Assessment of certainty of evidence

The certainty of the evidence for all outcomes will be judged using the Grading of Recommendations Assessment, Development and Evaluation working group methodology (GRADE Working Group) [31], across the domains of risk of bias, consistency, directness, precision and reporting bias. Certainty will be adjudicated as high, moderate, low or very low. For the main comparisons and outcomes, we will prepare Summary of Findings (SoF) tables [32],[29] and also interactive Summary of Findings (http://isof.epistemonikos.org/) tables. A SoF table with all the comparisons and outcomes will be presented as an appendix.

### Living evidence synthesis

An artificial intelligence algorithm deployed in the Coronavirus/COVID-19 topic of the L·OVE platform (https://app.iloveevidence.com/loves/5e6fdb9669c00e4ac072701d) will provide instant notification of articles with a high likelihood to be eligible. The authors will review these and will decide upon inclusion, and will update the [living web version](https://app.iloveevidence.com/living-reviews/5e7e748c53e53e78053563fd) of the review accordingly. We will consider resubmission to a journal if there is a change in the direction of the effect on the critical outcomes or a substantial modification to the certainty of the evidence.

This review is part of a larger project set up to produce multiple parallel systematic reviews relevant to COVID-19 [18].

## Data Availability

All data related to the project will be available. Epistemonikos Foundation will grant access to data.

## Acknowledgements

The members of the COVID-19 L·OVE Working Group and [Epistemonikos Foundation](https://www.epistemonikos.cl/) have made it possible to build the systems and compile the information needed by this project. Epistemonikos is a collaborative effort, based on the ongoing volunteer work of over a thousand contributors since 2012.

## Roles and contributions

GR conceived the protocol. GR and FV drafted the manuscript, and all other authors contributed to it. The corresponding author is the guarantor and declares that all authors meet authorship criteria and that no other authors meeting the criteria have been omitted.

The COVID-19 L·OVE Working Group was created by Epistemonikos and a number of expert teams in order to provide decision makers with the best evidence related to COVID-19. Up-to-date information about the group and its member organisations is available here: epistemonikos.cl/working-group

## PROSPERO registration

This protocol has been submitted (awaiting PROSPERO ID allocation).

## Ethics

As researchers will not access information that could lead to the identification of an individual participant, obtaining ethical approval was waived.

